# Wastewater Surveillance of SARS-CoV-2 across 40 U.S. states

**DOI:** 10.1101/2021.03.10.21253235

**Authors:** Fuqing Wu, Amy Xiao, Jianbo Zhang, Katya Moniz, Noriko Endo, Federica Armas, Mary Bushman, Peter R Chai, Claire Duvallet, Timothy B Erickson, Katelyn Foppe, Newsha Ghaeli, Xiaoqiong Gu, William P Hanage, Katherine H Huang, Wei Lin Lee, Mariana Matus, Kyle A McElroy, Steven F Rhode, Stefan Wuertz, Janelle Thompson, Eric J Alm

## Abstract

Wastewater-based disease surveillance is a promising approach for monitoring community outbreaks. Here we describe a nationwide campaign to monitor SARS-CoV-2 in the wastewater of 159 counties in 40 U.S. states, covering 13% of the U.S. population from February 18 to June 2, 2020. Out of 1,751 total samples analyzed, 846 samples were positive for SARS-CoV-2 RNA, with overall viral concentrations declining from April to May. Wastewater viral titers were consistent with, and appeared to precede, clinical COVID-19 surveillance indicators, including daily new cases. Wastewater surveillance had a high detection rate (>80%) of SARS-CoV-2 when the daily incidence exceeded 13 per 100,000 people. Detection rates were positively associated with wastewater treatment plant catchment size. To our knowledge, this work represents the largest-scale wastewater-based SARS-CoV-2 monitoring campaign to date, encompassing a wide diversity of wastewater treatment facilities and geographic locations. Our findings demonstrate that a national wastewater-based approach to disease surveillance may be feasible and effective.

## Introduction

COVID-19 was first reported in the United States on January 20, 2020, and spread to all 50 states and the District of Columbia by mid-March^1,2^. As of February 1, 2021, over 26 million confirmed cases and over 440,000 deaths have been reported in the U.S.^2^. Establishing a national COVID-19 surveillance system, like those for viral hepatitis and influenza, would be helpful for long-term monitoring of SARS-CoV-2, allowing healthcare officials to recognize and respond to new outbreaks efficiently and uniformly. However, COVID-19 poses specific challenges to clinical surveillance systems, with its long infectious incubation time (up to 14 days; median: 4-5 days) greatly increasing the risk of viral transmission and infection among the population before clinical reporting, contact tracing, and containment can occur^3,4^. “Test and trace” systems were rapidly overwhelmed in many countries early in the pandemic, and are often ineffective once a disease reaches exponential community spread^5,6^. The emergence of more infectious variants may exacerbate this problem^7,8^.

As a complementary approach to clinical disease surveillance, wastewater monitoring can help detect the presence of pathogens like the coronavirus SARS-CoV-2 across municipalities, and estimate disease incidence independent of individual testing^9–11^. Wastewater surveillance is less resource intensive than large-scale clinical testing, making it an optimal tool for unobtrusive, long-term monitoring as well as early identification of viral circulation in the population. Our recent findings^9,10^ along with work from other groups have described reliable detection of SARS-CoV-2 gene fragments in wastewater samples across the world, including Australia^12^, Brazil^13^, France, Netherlands^14^, Italy^15^, Spain^16^, and the U.S.^9,17^. Furthermore, longitudinal wastewater viral titers correlate with clinically diagnosed new COVID-19 cases, and trends in wastewater precede those in clinical reports by 4-10 days, suggesting that wastewater data could be used as an early warning of impending outbreaks to define public health and hospital planning^10^. The potential value of wastewater surveillance is gaining recognition, with the Centers for Disease Control and Prevention and several state and local health agencies initiating wastewater-based monitoring programs to supplement their COVID-19 responses^18^.

We implemented a nationwide COVID-19 surveillance campaign to measure viral concentrations of SARS-CoV-2 in the wastewater of 159 counties in 40 U.S. states, from February to June 2020. We investigated the detection rate and accuracy of wastewater surveillance of SARS-CoV-2 by comparing wastewater data to clinically reported case counts from state and local health agencies. We demonstrated the feasibility of utilizing wastewater surveillance as a supplement to national SARS-CoV-2 clinical reporting data to understand important past, current and future trends in viral dynamics.

## Results

We collected and processed 1,751 wastewater samples from 353 unique locations in 159 counties, in 40 U.S. states, from February 18 to June 2, 2020 (Fig. 1A). Of these samples, 1,687 were from locations that authorized the disclosure of their metadata. Individual samples represented catchments serving population sizes ranging from 65 to 5.3 million sewered individuals, with a median size of 31,745 people (Fig. S1a). In total, these wastewater samples covered 42.5 million people – approximately 13% of the U.S. population. To our knowledge, this is the largest dataset reporting temporal tracking of SARS-CoV-2 in wastewater. Samples were processed as they were received in the lab and quantified by real-time quantitative PCR (see Methods). 830 samples (49.1%) were positive for SARS-CoV-2 gene fragments.

**Figure 1.**
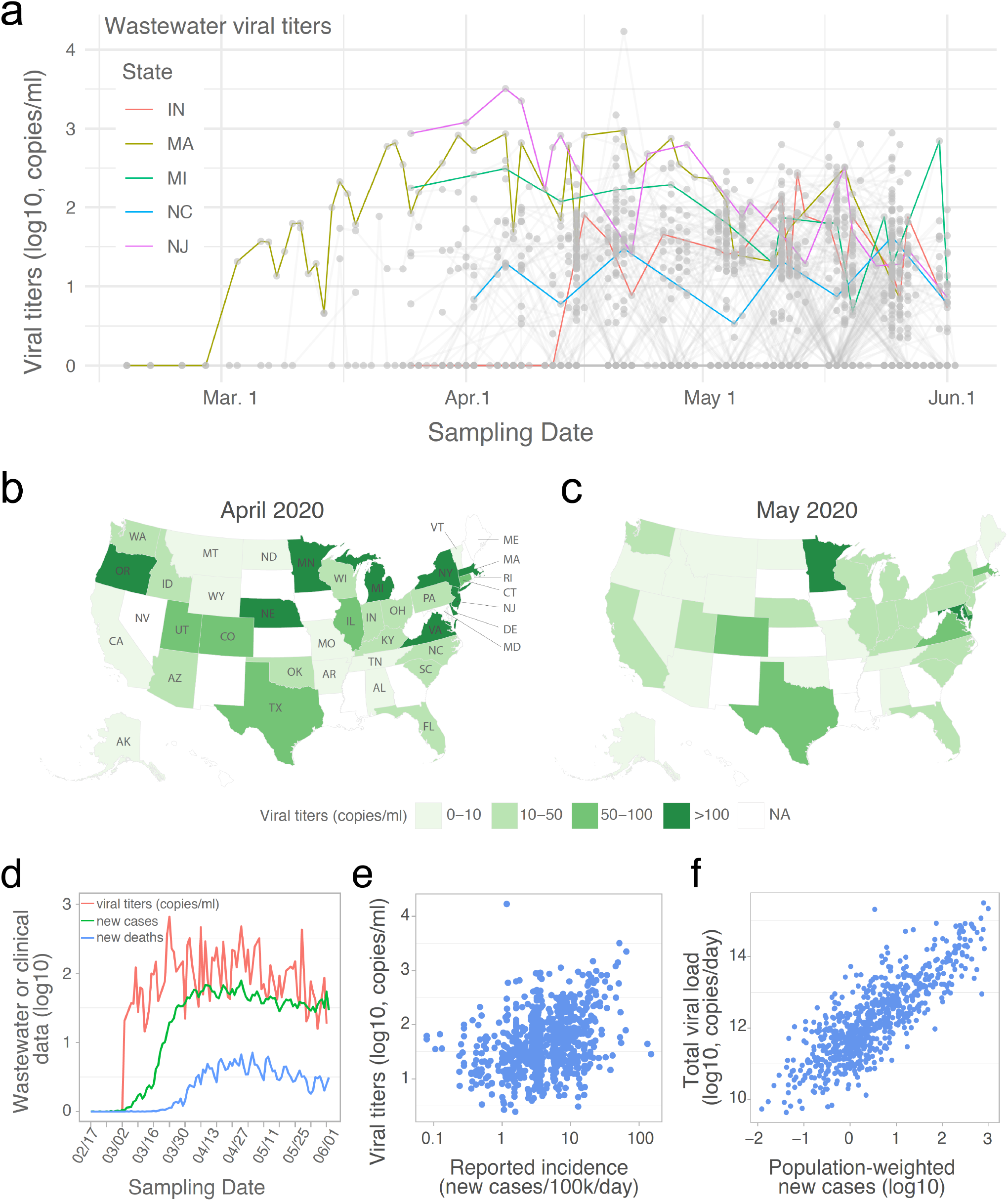
SARS-CoV-2 RNA gene copies in wastewater samples from 40 U.S. states. (a) Temporal profile of SARS-CoV-2 viral RNA gene copies (viral titers) in the wastewater samples collected from February 18 to June 2, 2020. Each grey point represents a sample, and the grey line connects samples collected from the same catchment. Temporal dynamics of mean viral titers from five U.S. states are highlighted. Negative samples (SARS-CoV-2 not detected) were assigned to ‘0’. (b-c) Mean viral titers for each state in April (b) and May (c). All the samples in April or May were aggregated by state. NA: data is not available for the state. (d) Temporal dynamics for the mean viral titers, daily new COVID-19 cases, and new deaths. Viral concentrations (red line) from positive wastewater samples were aggregated by date, and new cases (green line) and COVID-19-related new deaths (blue line) from the wastewater sample originated counties were also aggregated and averaged by date. (e) Association between viral titers in wastewater samples and the reported daily incidence rate in each sampled counties. (f) Association between the total viral load and estimated new cases in each of the catchment areas. Total viral load of SARS-CoV-2 in wastewater (copies/day) was calculated by multiplying SARS-CoV-2 concentration (copies/ml) by the daily average influent flow (ml/day) reported by the WWTP. Population weighted new cases was calculated as county new cases * catchment population / county population.

### Temporal dynamics of SARS-CoV-2 titers in wastewater samples from 159 counties in 40 states

For the month of March, we analyzed 86 wastewater samples from 25 counties (42 individual catchments), in 10 states. We observed significant heterogeneity in results at the county level. 44 of the 86 samples (51%) (from 12 counties in 8 states) were positive for SARS-CoV-2. In California, we found only 3 positive samples from the 14 sampling locations (21%) in the 7 counties sampled in March. On the other hand, we consistently detected SARS-CoV-2 in a Massachusetts wastewater treatment plant (WWTP) starting on March 3 (93% of samples), with viral titers increasing throughout the month (Fig. 1a). Positive samples were also found in more than two counties in Colorado, Oregon, and Texas.

In April, viral titers stopped increasing and became relatively stable for most sampling locations (Fig. 1a and Fig. S1). Of the samples tested in April, 52.9% (255 of 482) were SARS-CoV-2 positive: 173 samples from 44 counties had viral titers between 10-100 copies per ml of wastewater; 78 samples from 24 counties had viral titers higher than 100 copies per ml (Fig. 1a). We processed 1,092 samples from 154 counties in May. Of these, 69.5% (358/515) had positive SARS-CoV-2 titers of 10-100 copies per ml of wastewater, and 18.8% (97/515) had titers of >100 copies per ml (Fig. 1a). Analysis of April and May samples also showed the heterogeneity in viral titers at the county and catchment levels (Fig. S2).

We observed different dynamics in viral titers at the state level (Fig. 1a). Viral titers in New Jersey (NJ) were high in March samples, but started to decrease after April 8. Similar temporal dynamics were also observed in Michigan (MI), but with a smaller magnitude. Comparatively, viral titers in Indiana (IN) and North Carolina (NC) varied little over the sampling period (April and May). Nine states (Virginia, Delaware, Michigan, Minnesota, Massachusetts, Oregon, New York, Nebraska, and New Jersey) had titers higher than 100 copies per ml of wastewater in April (Fig. 1b). This number dropped to two states (Maryland and Minnesota) in May (Fig. 1c). Averaged across all states, the mean viral concentration was significantly higher in April than in May (Fig. S1b). Together, these data highlight that wastewater surveillance can be implemented to explore viral transmission at different geographic and temporal scales.

### Wastewater viral dynamics are consistent with clinical COVID-19 surveillance indicators

Next, we compared the wastewater viral titers with clinical surveillance data of COVID-19 across the U.S. Aggregating the positive wastewater data and daily new COVID-19 cases and new deaths by date, the mean viral titers increased from early March and became relatively stable between late March to late April, followed by a small downward trend until June 2 (Fig. 1d). This temporal profile mirrors the trends of clinical new cases and deaths at the national level, and precedes clinical data. Wastewater viral titers also reflected, and seemed to precede, the rise and fall of hospitalization and intensive care unit admissions (Fig. S3).

We also investigated the relationship between wastewater viral titers and daily COVID-19 incidence rates. Viral concentration in the wastewater is determined by the number of new infections, shedding rates from infected individuals, as well as total influent flow at the wastewater treatment plant^10^, which is linearly correlated with catchment population size (Fig. S4). A weak positive correlation was found between the incidence of daily new cases at the county level, and the wastewater viral titers at the catchment (wastewater treatment plant) level (Fig. 1e). We then compared total daily viral load for each catchment (viral concentrations detected at wastewater treatment plant, multiplied by the plant’s daily influent flow rate), and the estimated number of new cases in that catchment (county-level incidence rates multiplied by the catchment population). A linear relationship was observed between the total viral load and catchment size-normalized daily new cases (Fig. 1f), consistent with the hypothesis that average shedding rates are similar across catchments.

### Estimation of detection rate and accuracy of wastewater surveillance

Next, we investigated the detection rate of wastewater surveillance by comparing wastewater titers to new clinical cases on the sampling day (Methods). Using the reported daily incidence COVID-19 cases (i.e. daily new cases divided by the county population size), we calculated the percentage of positive wastewater samples for different incidence rates. Wastewater-based detections increased exponentially with the clinical incidence rate, reaching an 80% rate of detection at a clinical incidence of 13 cases per 100,000 people (Fig. 2a). For all positive wastewater samples at the county level, the associated incidence rates of daily new cases ranged from 0 – 149.6 cases per 100,000 people (median: 3.7 cases per 100,000 people) (Fig. 2b). In other words, wastewater-based surveillance was capable of detecting SARS-CoV-2 for one new reported case out of ∼27,000 people. However, this new case rate does not consider unreported infections in the population, which would lower the estimated detection limit.

**Figure 2.**
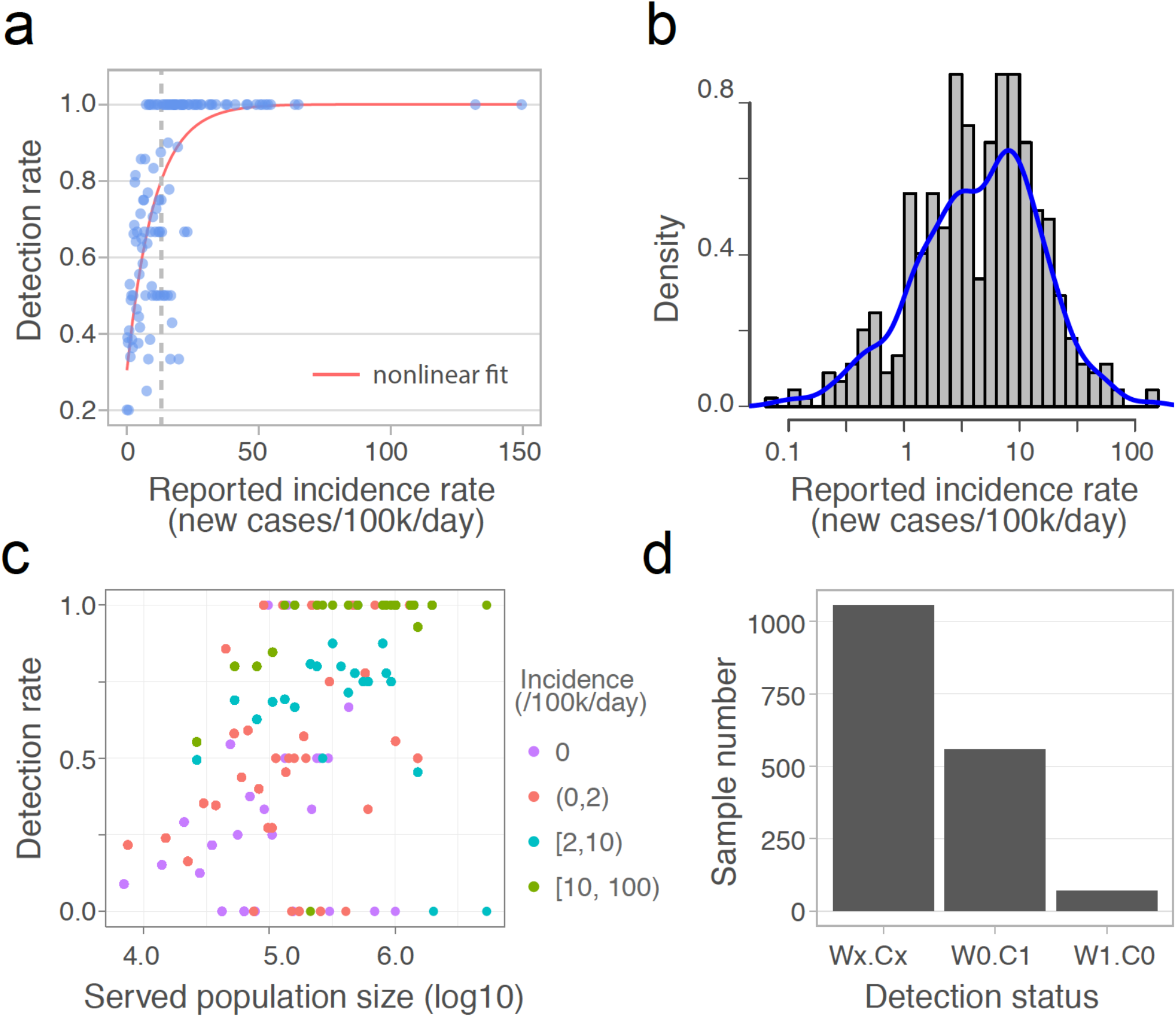
Detection rate and accuracy of wastewater-based SARS-CoV-2 surveillance. (a) Detection rate for varying daily incidence of COVID-19 cases. Each dot represents the percentage of positive wastewater samples for a constant incidence interval, and the red line is the nonlinear fit. The vertical dashed line indicates the incidence (x = 13) above which the fitted detection rate exceeds 0.8. (b) The distribution of daily incidence for the counties where SARS-CoV-2 was detected in the wastewater samples. Blue line is the Kernel density estimation of the daily incidence’s distribution. The median of the incidence is 3.7 cases per 100,000 people). (c) Relationship between the detection rate of positive wastewater samples from a given treatment plant and the population size served by that plant. Detection rate is binned by population size, and colored by the incidence intervals. (d) Detection status for all the samples (n = 1,687). Wx.Cx (x = 1 or 0): consistent results between wastewater data and clinical reports. W1.C1: SARS-CoV-2 detected in Wastewater (W1) and new Clinical cases reported (C1); W0.C0: no Wastewater detection (W0) and no new Clinical cases reported (C0); W0.C1: no Wastewater detection (W0) but new Clinical cases reported (C1); W1.C0: Wastewater detection (W1) but no new Clinical cases reported (C0).

To evaluate whether catchment size influences the probability of SARS-CoV-2 detection in wastewater samples, we analyzed the detection rate of positive samples from counties with equal daily incidence. As shown in Fig. 2c, detection rate is positively associated with the population size of wastewater treatment plant catchments for the majority of samples. 100% detection rates were disproportionately represented among samples with high incidence (>10 cases per 100,000 people) and large population sizes (>100,000 people). This result is consistent with our previous model simulations that the probability of SARS-CoV-2 detection in the wastewater increases with population size in communities with equal incidence^10^.

To evaluate the detection accuracy of wastewater surveillance, we compared the wastewater results with reported daily new clinical cases. For all 1,687 samples for which we had access to metadata, 1,057 (62.7%) exhibited results consistent with the geographically associated clinical data, meaning that SARS-CoV-2 was detected in the wastewater in areas with new clinical cases (759 samples, “W1.C1”), and not detected in areas where no new cases were reported (298 samples, “W0.C0”) (Fig. 2d). Of the remaining 630 samples, 559 had clinical cases but SARS-CoV-2 was not detected in the wastewater (“W0.C1”). Of these, 67.4% were from counties with incidence rates below the median of all wastewater samples (3.7 cases per 100,000). We also compared the pepper mild mottle virus levels (PMMoV), a stable and persistent indicator of fecal concentration in wastewater^9,19,20^, and found that PMMoV copies in the W0.C1 samples were slightly but significantly lower than in other samples (Fig. S5a), suggesting that sample dilution and low incidence rates may have contributed to wastewater non-detections. Finally, there were 71 samples (“W1.C0”) for which SARS-CoV-2 was detected, but there were no new clinical cases reported (Fig. 2d). Most of these samples’ viral titers ranged from 10 to 272 copies/ml (Fig.S5b). Comparison of the wastewater data against the 7-day averages of new clinical cases did not yield substantially different results (Fig. S5c-d).

## Discussion

In this study, we tested and quantified SARS-CoV-2 genome copies in 1,751 wastewater samples, collected from 159 U.S. counties in 40 states, using RT-qPCR. This nationwide campaign covered approximately 13% of the U.S. population and demonstrated that widespread wastewater surveillance is feasible and useful across various catchment sizes. Overall, viral titers increased starting in early March, and became relatively stable until April, followed by a small decrease in May. Thirty-eight out of 40 states we sampled issued stay-at-home orders or advisories between March 19 and April 7, and all 40 sampled states put statewide restrictions on activity in place between March 10 and April 6 (https://www.usatoday.com/storytelling/coronavirus-reopening-america-map/). These social distancing guidelines may have contributed to the relatively stable viral titers in April and downward trend in May.

Wastewater surveillance of SARS-CoV-2 has been widely employed in the U.S. and other countries, however, the detection rate and limit have been unclear. Our spatiotemporal wastewater dataset enabled us to address this question. Assuming an equal incidence rate throughout the county, our analysis showed that wastewater-based SARS-CoV-2 monitoring has a high chance (> 80%) of detecting the viral RNA when the incidence of daily new cases exceeds 13 cases per 100,000 people. Considering only positive wastewater samples, the median detection limit becomes 1 case per ∼27,000 people. Our analysis is based on case reports from public health agencies, which are likely underestimates of true infection rates.

This study has several limitations. First, all surveillance data are limited by sampling regimes which could introduce bias through either the frequency of sampling or the specific locations sampled, which is true for both wastewater surveillance and case counts reported by public health authorities. A more unbiased sampling strategy including representative locations and appropriate time intervals would improve our estimate about the viral transmission in the population. Second, sample numbers were biased by month, 5.1% samples were from March and 55.5% samples were from May, thus our analysis may be affected by the low sampling resolution during the early stage of the pandemic. Third, all the samples were collected by each wastewater treatment plant, and mailed to us for analysis, and thus variation in sample collection and transport conditions may have further influenced data comparability. National implementation of a wastewater-based detection system would require standard operating procedures for sample collection, local processing, and analysis.

Here, we reported wastewater levels of SARS-CoV-2 in 159 counties in 40 U.S. states, from mid-February to early June 2020, and showed that wastewater data largely parallels and precedes clinical and epidemiological indicators of COVID-19 pandemics. Across the country, wastewater surveillance had a high SARS-CoV-2 detection rate (>80%) when the local daily incidence exceeded 13 reported cases per 100,000 people. The detection rate was positively associated with catchment population size. To our knowledge, this is the largest nationwide investigation of SARS-CoV-2 in wastewater samples in the U.S. during the COVID-19 pandemic, with samples from 353 catchments representing 13% of the U.S. population. This noninvasive and cost-effective approach could be employed as a complementary tool for long-term monitoring of SARS-CoV-2 – as well as other infectious diseases and other health-relevant biomarkers - across the United States.

## Methods

### Sample collection and viral inactivation

We initiated a national call for wastewater samples to quantify SARS-CoV-2 viral load in wastewater catchment areas from mid-February to early June 2020. Eligible sites included public works and wastewater authorities across the continental United States. Samples were collected by each wastewater treatment facility on a voluntary basis. Raw wastewater samples were collected from the wastewater treatment plants or catchments in 40 U.S. states and stored at 4C before being mailed to the laboratory for analysis. Samples were processed using the method as previously described^9,10^. Briefly, samples were first inactivated by ultraviolet light for 20 mins and heat treatment in a 60C water bath for 90 mins. Pasteurized samples were vacuum filtered with a 0.22-um polyethersulfone membrane to remove cell debris and solid materials. Supernatant was stored at 4C, and used for the viral enrichment.

### Viral precipitation, RNA extraction, reverse transcription and quantitative PCR (RT-qPCR)

We processed the samples starting from viral enrichment to quantitative PCR with two comparable methods as previously described^10^. 60 samples from deer island wastewater treatment plant were processed with both two methods, and no significant difference between viral titers was observed^10^. 1023 samples were processed with Method I, and 729 samples were processed with Method II. Briefly, viral particles in 40-ml filtrate were precipitated with polyethylene glycol 8000 (10% w/v, Millipore sigma) and NaCl (0.3M, Millipore sigma) in Method I. Viral pellet was resuspended in 1.5 mL Trizol reagent (Cat# 15596026, Thermo Fisher Scientific) for RNA extraction. cDNA was synthesized by reverse transcription (RT) based on the manufacturer’s protocol (M0368, New England Biosciences), followed by real-time PCR with the TaqMan® Fast Advanced Master Mix and U.S. CDC N1, N2 primer/probes. The qPCR reaction was carried out for 48 cycles using Bio-Rad CFX96 Real-Time PCR Detection System with following program: polymerase activation (95°C for 2 min), PCR (48 cycles, denature at 95°C for 1 s, and anneal/extend at 55°C for 30 s). 15 ml of filtrate in Method II were first concentrated with 10 kDa Amicon Ultra Centrifugal Filter (Sigma, Cat# UFC9010) to 150 ∼ 200 ul, which is further lysed with 600 ul AVL buffer (Qiagen, Cat# 19073) for RNA extraction (Qiagen RNeasy kit, Cat# 74182). The eluted RNA was used for one-step RT-PCR with TaqMan™ Fast Virus 1-Step Master Mix (Thermofisher, Cat# 4444436), based on the following protocol: 50°C 10 mins for reverse transcription, 95°C 20 s for RT inactivation and initial denaturation, and 48 cycles of denature (95°C 1 s) and anneal/extend (55°C 30 s).

Ct values for N1 or N2 primer sets were first converted to viral gene copies in the cDNA sample (copies per ul of cDNA) based on the standard curves established with the positive control plasmid (Method I) or Twist SARS-CoV-2 RNA (Method II) (*10*). The concentration was further converted to viral gene copies per microliter of the wastewater sample by multiplying the dilution factor. For Method I, the dilution factor is: the volume of total cDNA * the total volume of RNA / (the volume of RNA used for reverse transcription * the starting volume of filtered wastewater sample). For Method II, the dilution factor is: The total volume of RNA / the starting volume of filtered wastewater sample). Two or three replicates were performed for each primer set, averaged within each primer set and then across primers to derive the concentration values. We also measured PMMoV concentration in the sample as an internal reference for wastewater samples. We performed two technical replicates for each sample and converted the mean Ct values to relative concentrations of viral particles based on the standard curve^9^ and sample’s dilution factor.

### Clinical data collection and detection rate analysis

County-level clinical data including cumulative COVID-19 cases and deaths were downloaded from USAFACTS (https://usafacts.org/visualizations/coronavirus-covid-19-spread-map/). Daily new cases or deaths were generated through using the cumulative data on one day to subtract the data before that day. We compared wastewater viral titers to clinical data reported for the day on which the sample was obtained. For the detection accuracy analysis in Fig. S5c-d, we also compared the wastewater data against the 7-day moving average of new clinical cases (current day + 6 preceding days / 7). Hospitalizations and positive rates of testing for each state were downloaded from The COVID Tracking Project (https://covidtracking.com/).

Incidence rate of daily new cases was calculated using reported daily new cases in the county divided by the county population size. In Fig. 2a, we computed the detection rates, percentage of positive wastewater samples, for a constant interval (0.2 cases per 100,000 people) of daily incidence, starting from 0 to 149.6 cases per 100,000 people (maximum daily incidence). The results were fitted using an exponential decay function with formula: y ∼ *k*_1_ + *V*_max_ * (*k*_2_ - exp(-x / *tau*), starting from *V_max_* = 10, tau = 1, and *k*_1_ = 0.2, *k*_2_ = 1.15. To estimate the distribution of daily incidence for all the positive samples, we first aggregated wastewater viral titers for each county, since clinical cases were reported at the county level. Then we selected the positive samples and plotted the histogram and Kernel density estimation of the distribution of daily incidence (Fig. 2b). In Fig. 2c, we investigated the relationship between detection rate of positive wastewater samples and the catchment population size. We first separated the samples based on four different daily incidences (0, (0,2), [2,10), and [10,100), per 100,000 people) in the county where the sample was obtained. For each daily incidence, we computed the detection rate for a constant interval of population size, i.e. the maximum minus minimum of population size, and divided by the number of bins (n=200). All the analysis was done with R (3.5.0).

## Data Availability

The data presented in the figures, and the code for the analysis, and that support the other findings of this study are available from the corresponding author on reasonable request.

## Acknowledgments

We thank the management and sampling teams at all the wastewater treatment facilities who worked with us in providing the samples for analysis. We thank Penny Chisholm (MIT) and Allison Coe (MIT) for access to equipment and other supplies, Shandrina Burns (MIT) for logistical support. Finally, we express our deep gratitude to all healthcare professionals and first-line responders who have been caring for patients with COVID-19.

This work was supported by the Center for Microbiome Informatics and Therapeutics and Intra-CREATE Thematic Grant (Cities) grant NRF2019-THE001-0003a to JT and EJA; National Institute on Drug Abuse of the National Institutes of Health award numbers K23DA044874; and R44DA051106 to MM and PRC, Hans and Mavis Psychosocial Foundation funding, and e-ink corporation funding to PRC; funding from the Morris-Singer Foundation and NIH award R01AI106786 to WPH; funds from the Massachusetts Consortium on Pathogen Readiness and China Evergrande Group to TBE, PRC, MM, and EJA; funding from the Singapore Ministry of Education and National Research Foundation through an RCE award to Singapore Centre for Environmental Life Sciences Engineering (SCELSE) to SW and JT. The content is solely the responsibility of the authors and does not necessarily represent the official views of the funding institutions.

## Supplemental figures

**Figure S1.**
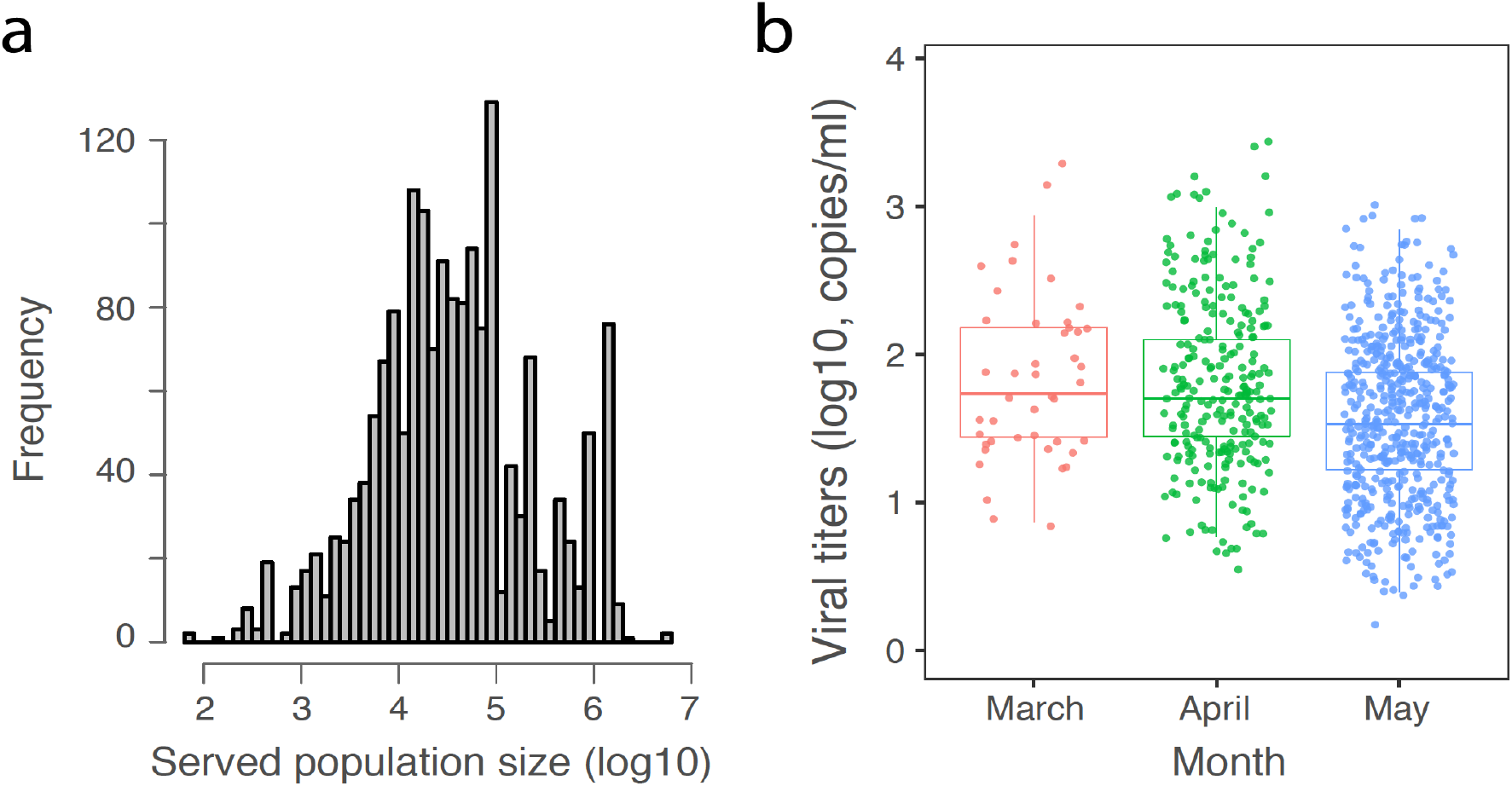
Distribution of the wastewater treatment plant serving population size, and viral titers in positive wastewater samples by month. (a) Histogram of population sizes served by the sampled wastewater treatment plants. The median served population size is 31,745. (b) Viral titers in positive wastewater samples by month. The box represents the interquartile range of viral titers for each month, the horizontal line inside the box is the median. Significant difference was found between the mean viral titers in April and May (Welch’s t-test, p-value = 0.025).

**Figure S2.**
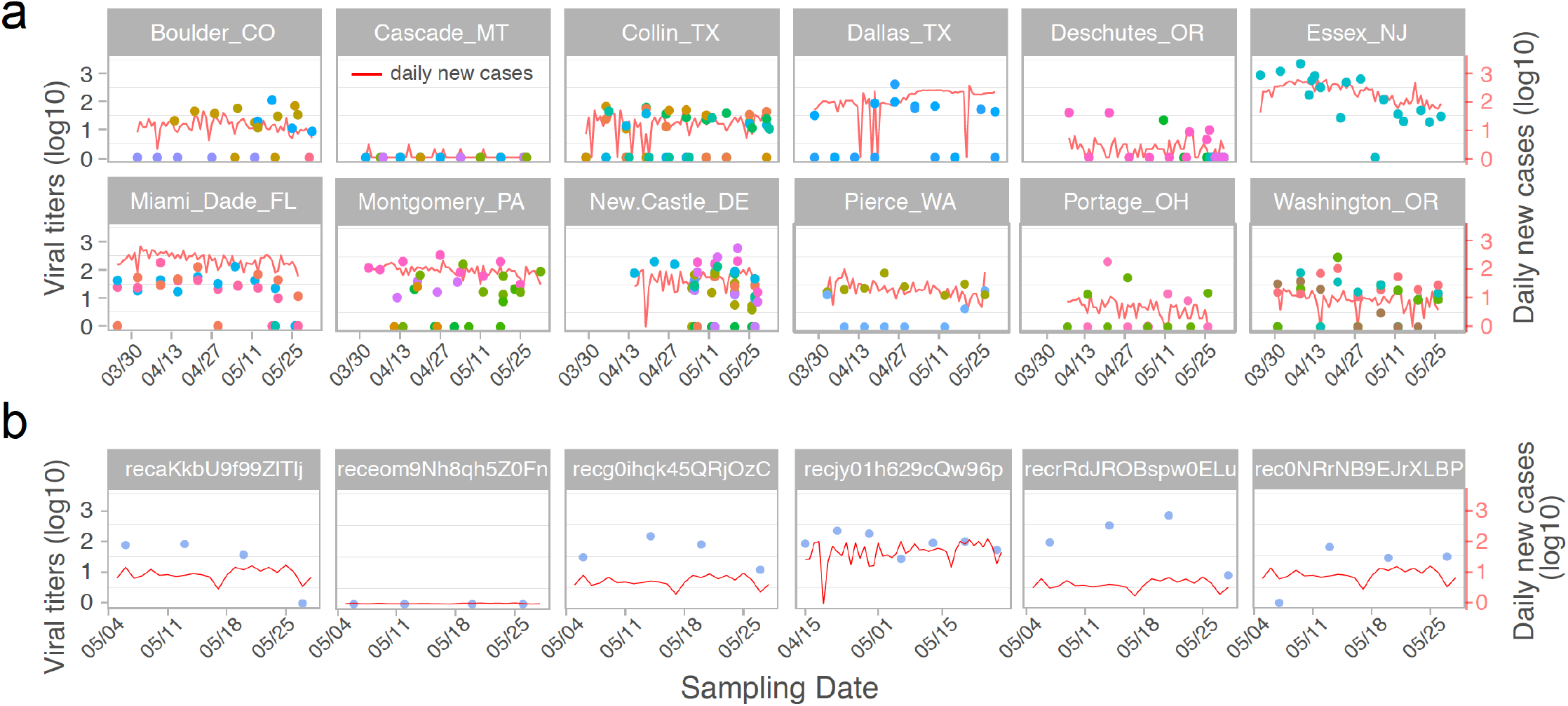
Temporal SARS-CoV-2 titers at the county level and catchment level. (a) Temporal viral titers in 12 representative counties. Each dot is a sample, and colored by the sampling catchments in the county. Red lines represent the daily new cases in the corresponding counties during the sampling period. (b) Temporal viral titers for samples collected in six different locations in the New Castle County of Delaware. Red lines represent the weighted daily new cases (daily new cases in the county multiplying the catchment served population size and divided by the county population size) during the sampling period.

**Figure S3.**
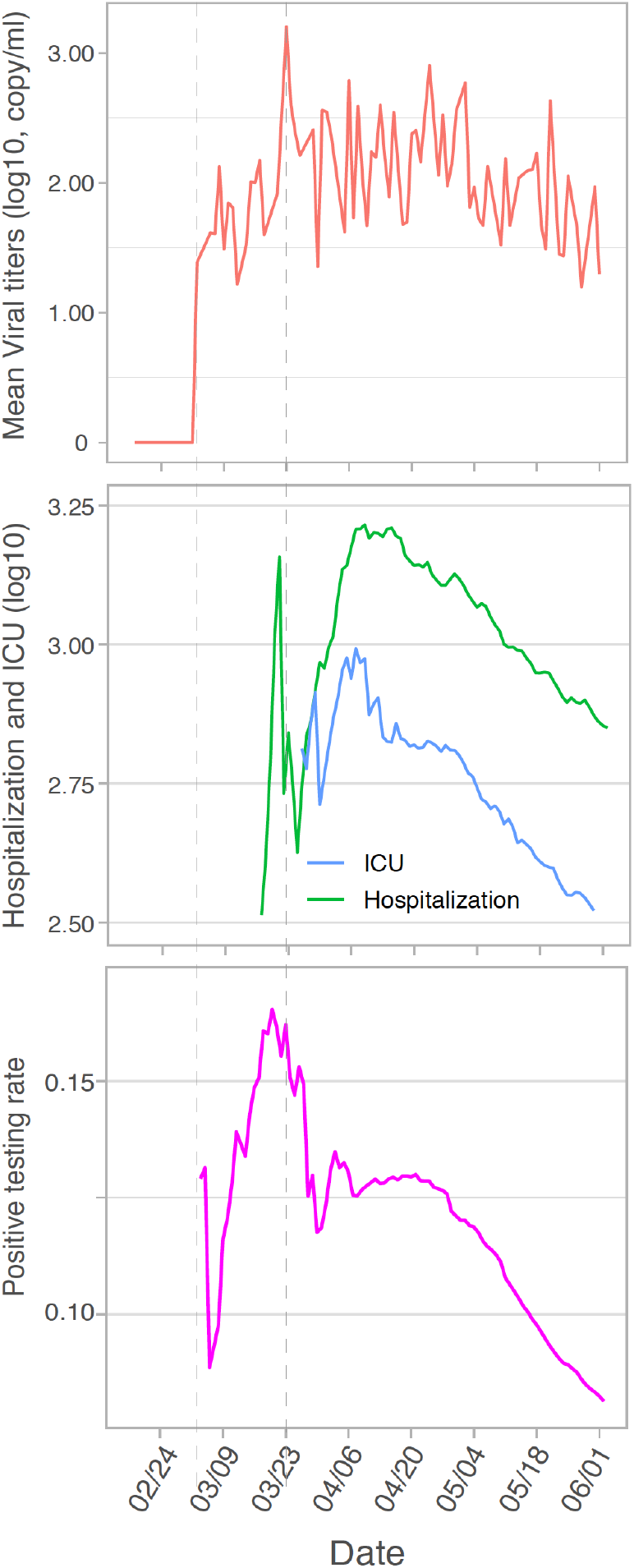
Wastewater viral titers (top) and clinical COVID-19 surveillance indicators including hospitalization and intensive care unit admissions (middle), and the testing positive rates (bottom) from February to June. Positive wastewater data from all the sampling locations were aggregated by date using the mean function. Clinical data from the 40 sampled states were aggregated in the same way.

**Figure S4.**
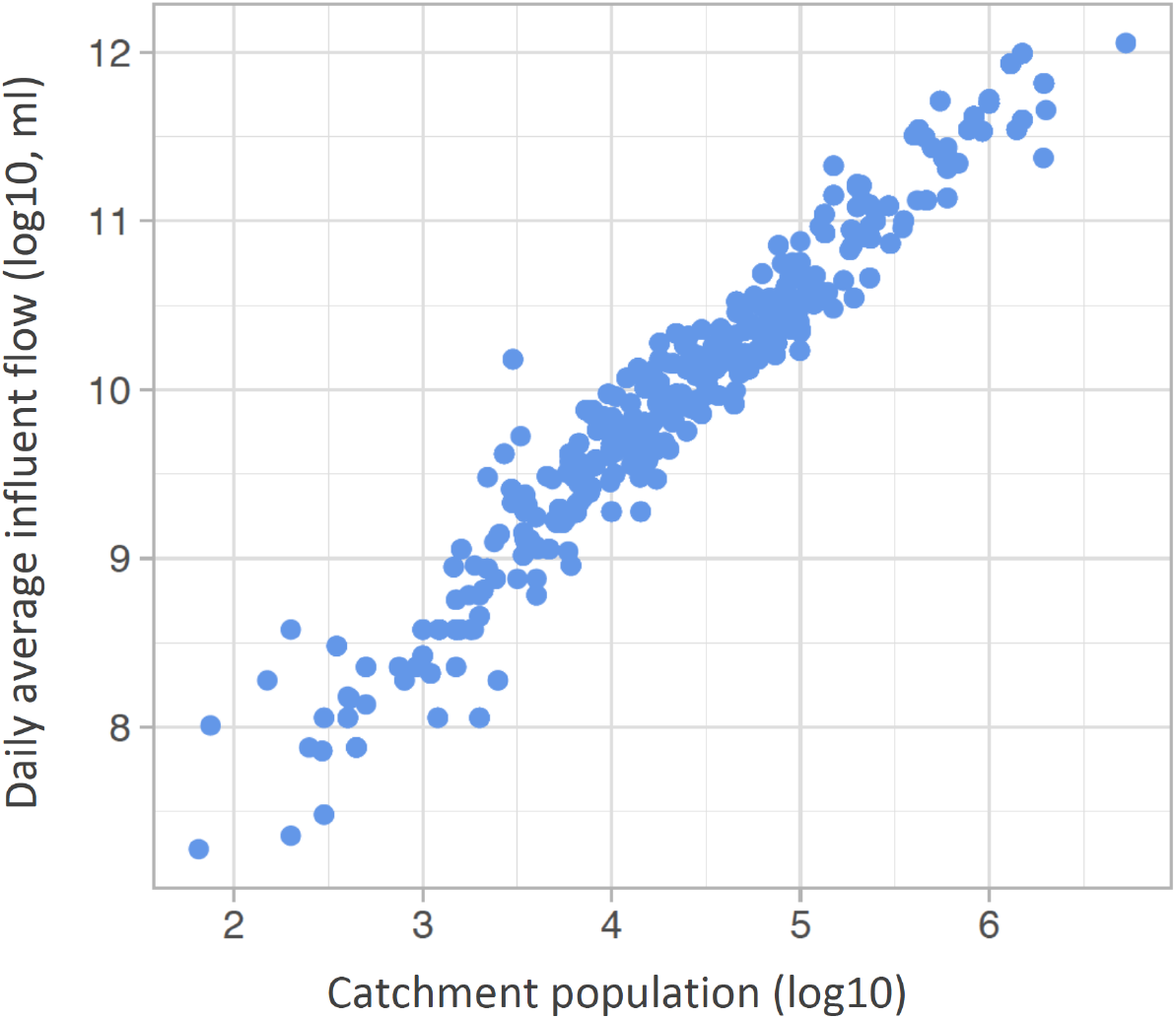
Daily average influent flow at the wastewater treatment plant is correlated with catchment population size.

**Figure S5.**
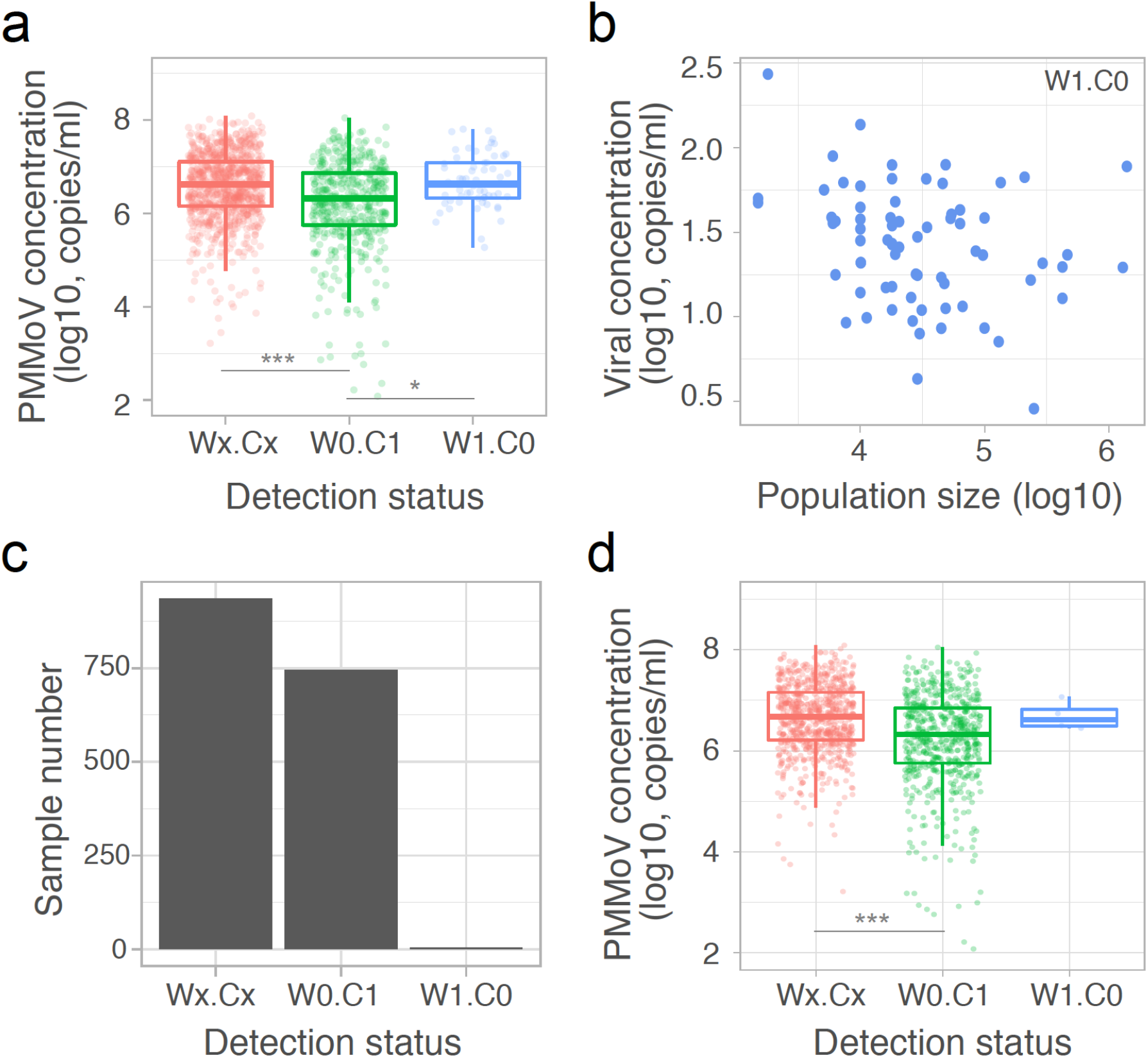
PMMoV concentrations in wastewater samples, and detection status with 7-day averages of new COVID-19 cases. (a) PMMoV concentrations in the Wx.Cx (x = 0 or 1), W0.C1, and W1.C0 groups. W1.C1: SARS-CoV-2 detected in Wastewater and new Clinical cases reported; W0.C0: no Wastewater detection and no new Clinical cases reported; W0.C1: no Wastewater detection but new Clinical cases reported; W1.C0: Wastewater detection but no new Clinical cases reported. Significant differences of PMMoV concentrations were found between W0.C1 and Wx.Cx or W1.C0 groups (Welch’s t-test, and symbol *: p-value < 0.05; ***: p-value < 0.001). (b) Viral titers and the served population size for the W1.C0 samples. Most of these samples were from catchments serving 10,000 ∼ 100,000 population, with viral titers ranging from 10 to 100 copies per ml of wastewater. (c-d) Detection status for wastewater data against 7-day averages of new clinical cases (c), and PMMoV concentrations in the Wx.Cx, W0.C1, and W1.C0 groups (d). Significant difference was found between Wx.Cx and W0.C1 groups (Welch’s t-test, and symbol ***: p-value < 0.001).

